# A Non-targeted Proteomics Newborn Screening Platform for Genetic Disorders

**DOI:** 10.1101/2024.01.23.24301545

**Authors:** Hirofumi Shibata, Daisuke Nakajima, Ryo Konno, Atsuhi Hijikata, Motoko Higashiguchi, Hiroshi Nihira, Saeko Shimodera, Takayuki Miyamoto, Masahiko Nishitani-Isa, Eitaro Hiejima, Kazushi Izawa, Junko Takita, Toshio Heike, Ken Okamura, Hidenori Ohnishi, Masataka Ishimura, Satoshi Okada, Motoi Yamashita, Tomohiro Morio, Hirokazu Kanegane, Kohsuke Imai, Yasuko Nakamura, Shigeaki Nonoyama, Toru Uchiyama, Masafumi Onodera, Ryuta Nishikomori, Osamu Ohara, Yusuke Kawashima, Takahiro Yasumi

**Affiliations:** Department of Pediatrics, Kyoto University Graduate School of Medicine, Kyoto, Japan; Department of Applied Genomics, Kazusa DNA Research Institute, Kisarazu, Japan; School of Life Sciences, Tokyo University of Pharmacy and Life Sciences, Tokyo, Japan; Department of Dermatology, Faculty of Medicine, Yamagata University, Yamagata, Japan; Department of Pediatrics, Gifu University Graduate School of Medicine, Gifu, Japan; Department of Pediatrics, Graduate School of Medical Sciences, Kyushu University, Fukuoka, Japan; Department of Pediatrics, Hiroshima University Graduate School of Biomedical and Health Sciences, Hiroshima, Japan; Department of Pediatrics and Developmental Biology, Graduate School of Medical and Dental Sciences, Tokyo Medical and Dental University (TMDU), Tokyo, Japan; Department of Child Health and Development, Graduate School of Medical and Dental Sciences, Tokyo Medical and Dental University (TMDU), Tokyo, Japan; Department of Pediatrics, National Defense Medical College, Tokorozawa, Japan; Department of Human Genetics, National Center for Child Health and Development, Tokyo, Japan; Gene & Cell Therapy Promotion Center, National Center for Child Health and Development, Tokyo, Japan; Department of Pediatrics and Child Health, Kurume University School of Medicine, Kurume, Japan

## Abstract

Newborn screening using dried blood spot (DBS) samples has made a substantial contribution to public healthcare by detecting patients with genetic disorders as neonates. Targeted measurements of nucleic acids and metabolites have played major roles in newborn screening to date, while the feasibility of new non-targeted approaches, including genome-wide DNA sequencing, has been explored. Here, we investigated the applicability of non-targeted quantitative proteomics analysis to newborn screening for genetic diseases. DBS protein profiling allowed monitoring of levels of proteins encoded by 2912 genes, including 1106 listed in the Online Mendelian Inheritance in Man database, in healthy newborn samples, and was useful in identifying patients with inborn errors of immunity by detecting reduced levels of disease causative proteins and cell-phenotypical alterations. Our results indicate that application of non-targeted quantitative protein profiling of DBS samples can forge a new path in screening for genetic disorders.

## Introduction

Dried blood spot (DBS) samples, comprising spotted blood dried in filter paper, are widely used for pre-symptomatic disease screening because of their simplicity in terms of sampling, shipping, and storage. Since Dr. Guthrie introduced the first screening test for phenylketonuria (GUTHRIE and SUSI, 1963), newborn screening (NBS) programs using DBS samples have made considerable contributions to public healthcare. NBS can save neonates with serious inborn disorders from developing devastating symptoms, by introducing appropriate therapeutic intervention before disease onset, which can improve their long-term prognosis. Notably, the emergence of tandem mass spectrometry has enabled use of a single platform to simultaneously screen for a wide range of inborn errors of metabolism (IEMs), playing a critical role in the spread of NBS (Wilcken et al., 2003), and serving as a successful example of the power of non-targeted measurement platforms for pre-symptomatic screening for inherited diseases, followed by confirmatory diagnosis by genetic analyses.

Although NBS for IEMs can be achieved by assessing metabolic function through monitoring activity of metabolites or metabolic enzymes, direct measurement of molecular function is frequently too complex and infeasible in the context of NBS for other inherited conditions. As recent advances in DNA sequencing technology have dramatically improved cost-performance and throughput, the use of genome-wide sequencing has been proposed as a new alternative modality for NBS (Berg et al., 2017); however, there remain many technical, clinical, ethical, and social issues to be addressed before implementation of genome sequencing for NBS (Spiekerkoetter et al., 2023; Ulph and Bennett, 2023). In addition, concerns regarding the use of genome sequencing persist, due to an inability to predict gene product functionality from structural genetic information with high accuracy; in the absence of accumulated in-depth genotype-phenotype data, gene structure information cannot be linked with clinical phenotypes. Indeed, Adhikari et al. reported that exome sequencing was not sufficiently sensitive or specific as a primary screen for use in NBS for IEM, while it is useful as a second-tier test for some infants who test positive in first-tier screening, assisted by tandem mass spectrometry (Adhikari et al., 2020).

As the majority of inherited diseases occur due to loss or gain of protein function, we hypothesized that measurement of the quality and/or quantity of proteins encoded by causative genes could represent a new modality for NBS with advantages over structural gene analyses. We recently reported that proteome analysis of peripheral blood mononuclear cells can complement genetic analysis in identifying the causes of inborn errors of immunity (IEIs) in undiagnosed patients (Sakura et al., 2023). Here, we explore whether proteome analysis of DBS samples can contribute to the diagnosis/screening of IEIs and other genetic disorders. Targeted quantitative measurement of proteins of interest in DBS have been reported as test trials (Collins et al., 2020; Dezfouli et al., 2020); however, until recently, cost and throughput issues have not been surmounted. Recent advances in proteome technology have allowed us to address these issues; for example, advanced liquid chromatography-assisted mass spectrometry (LC-MS/MS) in data-independent acquisition (DIA) mode facilitates highly sensitive quantification of protein levels (Kawashima et al., 2022; Kawashima et al., 2019). Furthermore, a non-targeted method for in-depth DBS protein profiling has also been developed (Nakajima et al., 2020). These technical advances have allowed us to overcome the obstacles impeding progress in proteome analysis-based NBS.

In this context, the aims of the present study were to evaluate whether non-targeted proteomics analysis of newborn DBS by LC-MS/MS could be used to identify and quantify disease-causing or -related proteins, thereby clarifying the molecular phenotypes underlying genetic disorders at a pre-symptomatic stage. For this purpose, DBS samples from 40 healthy newborns were first subjected to non-targeted proteomics analysis, to assess their consistency and reliability for use in protein profiling by comparison with similar data generated from healthy adult samples. Subsequently, DBS samples from patients with IEIs, as a representative disease spectrum primarily caused by defects in genes expressed in blood, were analyzed to determine whether patients with these conditions can be identified by quantitative protein profiling. Our results indicate that non-targeted DBS proteomics can sensitively detect changes in molecular and cellular phenotypes underlying many IEIs, indicating that this approach may be useful for newborn diagnosis and/or screening for genetic diseases.

## Results

### Comparison of protein profiles from healthy newborn and healthy adult samples

LC-MS/MS and non-targeted deep proteome analyses of DBS samples from 40 healthy newborns enabled the identification and quantification of 3587 proteins within the identification threshold (Table S1, https://doi.org/10.5281/zenodo.10539971). These data were filtered to include only proteins with ≥ 2 identified peptides, resulting in a total of 2912 proteins that could be quantified within measurable ranges in all newborn samples. Of these, 1106 proteins encoded by genes listed in the Online Mendelian Inheritance in Man (OMIM) database were subjected to analysis after standardization based on β-actin levels in the same sample (Table S2, https://doi.org/10.5281/zenodo.10539971). We also analyzed samples from eight healthy adults. Correlation analysis indicated that samples in the newborn and adult DBS groups showed high within-group coefficient values (Pearson r = 0.849–0.973 and 0.898–0.962, respectively). Next, we conducted multiple evaluation of Pearson’s correlation coefficient values between the two sample types, along with principal component analysis, and the results showed that newborn samples clustered separately from adult samples (Fig. 1, A and B). Detailed analysis of protein levels to search for differentially expressed proteins (DEPs) identified 954 upregulated and 108 downregulated proteins in newborn, relative to adult, samples (Fig. 2 A). Upregulated proteins included components of fetal and embryonic hemoglobin (*HBG1/HBG2* and *HBE1/HBZ*, respectively) and alfa fetoprotein (*AFP*). Downregulated proteins included antithrombin III (*SERPINC1*), heparin cofactor 2 (*SERPIND1*), protein C (*PROC*), haptoglobin (*HP*), Hb subunit delta (*HBD*), immunoglobulin heavy constant mu (*IGHM*), and joining chain of multimeric IgA and IgM (*JCHAIN*) (Fig. 2, B and C, and Fig. S1). In Gene Ontology (GO) analysis of DEPs, translational initiation and regulation of complement activation pathways were ranked as highly enriched for proteins upregulated and downregulated in newborn relative to adult samples, respectively (Fig. 2 D and E). These results are generally consistent with previous reports (Kanakoudi et al., 1995; Reverdiau-Moalic et al., 1996) and collectively indicate the reliability of the current method for non-targeted quantitative evaluation of proteins in DBS samples.

**Figure 1.**
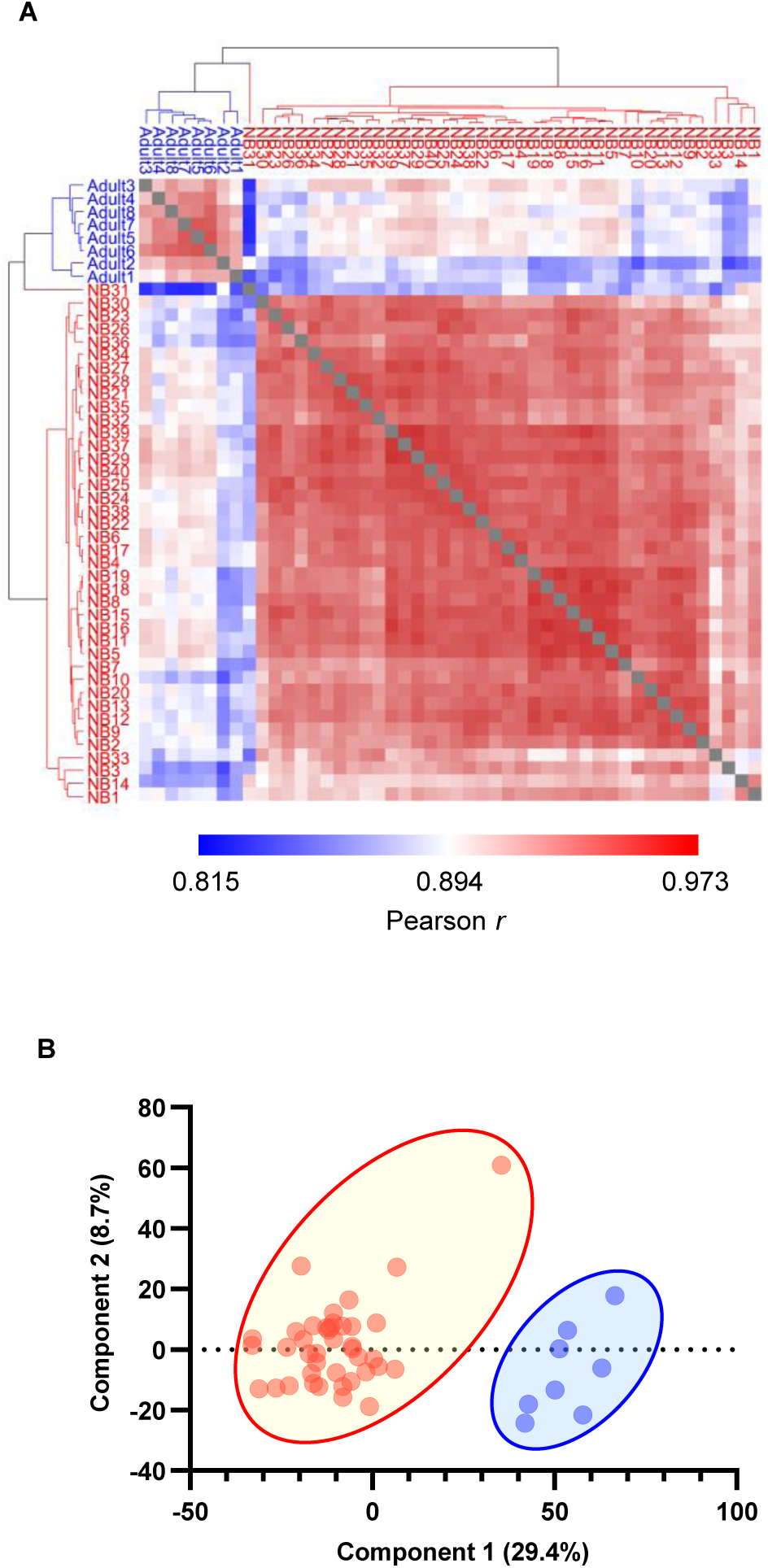
Proteomics analysis distinguishes healthy newborn dried blood spot (DBS) samples from those of adults. **(A)** Clustered heatmap of Pearson correlation coefficient values between all healthy newborn and adult DBS sample pairs. Dark red denotes higher correlation and dark blue denotes lower correlation (Pearson r = 0.82–0.96). **(B)** Principal component analysis of healthy newborn and adult DBS sample pairs. Red denotes newborn (NB) and blue denotes adult DBS samples.

**Figure 2.**
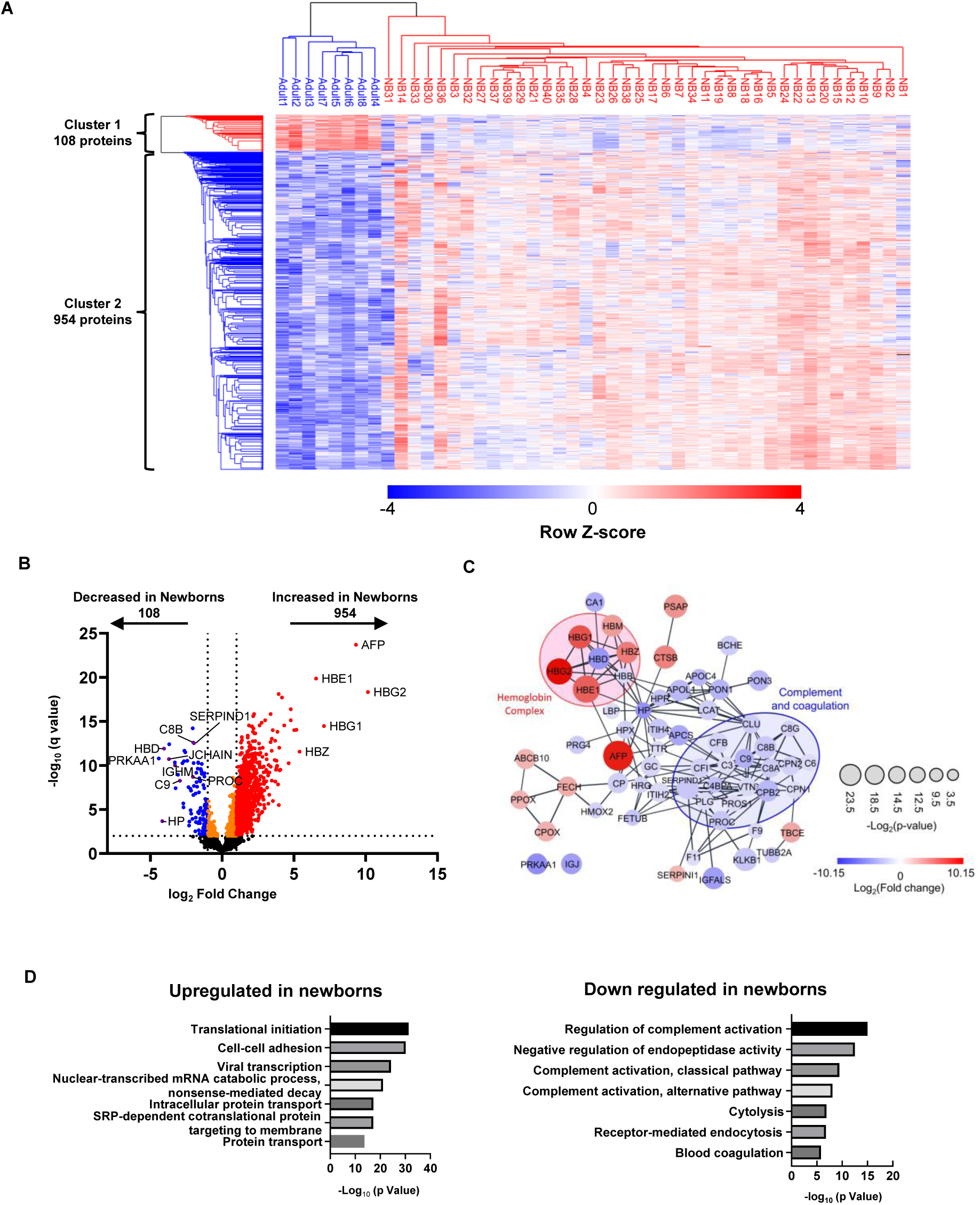
Proteins differentially expressed between healthy newborn and adult samples. **(A)** Heatmap of hierarchical clustering of all differentially expressed proteins (DEPs) detected by comparison between healthy newborn and adult samples. Red, upregulated DEPs; blue, downregulated DEPs. **(B)** Volcano plot showing DEPs in dried blood spot samples from healthy newborns compared with those from healthy adults. **(C)** Protein-protein interaction network analysis illustrating the top 100 upregulated and downregulated proteins (q-value < 0.05), contrasting healthy newborns and adults; network generated in Cytoscape using the STRING database. Node size corresponds to P value, while colors (red and blue) signify DEPs; red and blue circles represent the ‘Hemoglobin complex’ and ‘Complement and coagulation’ Gene Ontology (GO) terms, respectively. **(D)** GO enrichment analysis of DEPs. Bar charts showing the top 10 GO terms for biological processes upregulated (left) and downregulated (right) in newborn samples relative to adult samples.

### Proteins causative of various genetic disorders can be quantified in neonatal DBS samples

Next, we considered which diseases are feasible potential targets for diagnosis/screening by DIA-LC-MS/MS using DBS samples. Diseases with autosomal recessive or X-linked inheritance were identified as primary targets; as diseases with autosomal dominant inheritance are caused by anomaly of a single allele, while the other allele remains intact, which would be expected to minimally affect protein levels, they were not considered ideal for screening using this approach. Of the 1110 identified proteins listed in OMIM, 781 were related to diseases with autosomal recessive or X-linked inheritance, of which, 432, 71, and 139 proteins were categorized as metabolic/neurological, hematological, and immunological diseases, respectively (Table S2 and Fig. S2 A). Of the 139 proteins related to immunological diseases, 124 were included as causative molecules in the latest list of IEIs (Table S2) and most of these were distributed similarly in healthy newborn and adult samples (Fig. S2 B). To explore the possibility of direct detection of changes in protein levels caused by genetic mutations and their use for diagnosis/screening of genetic disorders, we next focused our efforts on characterization of DBS protein levels in samples from patients with IEIs (Table 1), as most IEI causative proteins are abundantly expressed in peripheral blood.

**Table 1.**
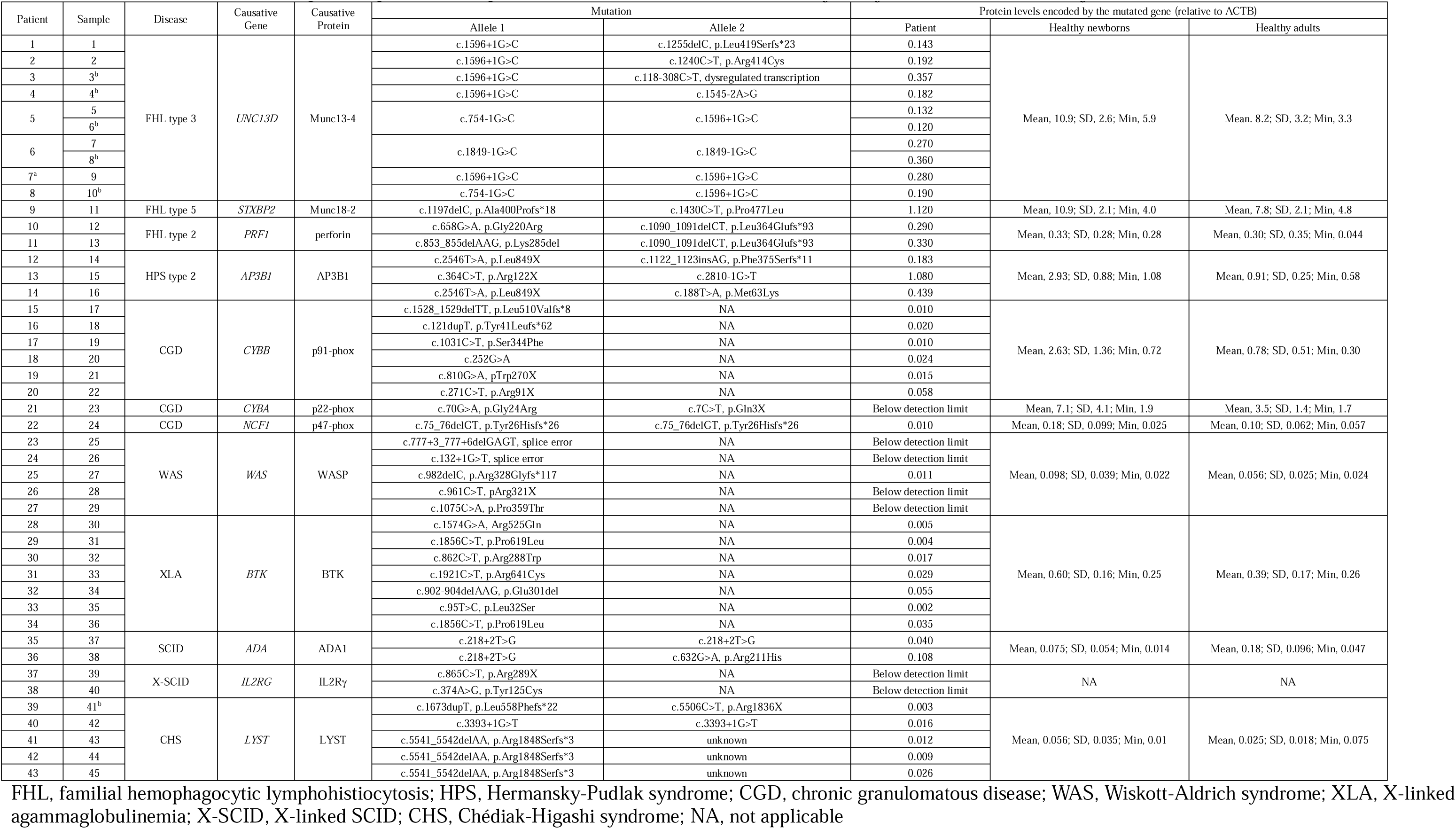
Details of 45 dried blood spot samples from 43 patients with inborn errors of immunity analyzed in the current study.

### Primary effect of IEI gene mutations on levels of their encoded proteins

First, we evaluated whether levels of IEI disease-causing proteins were exclusively reduced in patient samples and suitable for use in disease diagnosis. All 10 samples from patients with familial hemophagocytic lymphohistiocytosis (FHL) type 3, including five obtained before and five obtained after the onset of disease, showed significant reductions in Munc13-4 levels compared to control samples (Fig. 3 A and Fig. S3). This finding is consistent with reports showing that FHL type 3 can be reliably diagnosed by assessing intra-platelet Munc13-4 levels (Murata et al., 2011; Shibata et al., 2018). A similar result was observed in a patient with FHL type 5, in that Munc18-2 levels were markedly lower in the patient than in control samples (Fig. 3 B). By contrast, perforin levels did not differ significantly between patients with FHL type 2 and controls (Fig. 3 D), although flow cytometry showed that perforin levels were reduced in natural killer cells from patients (Fig. S4). Analysis of protein profiles in patients with other IEIs showed that levels of p91-phox, Wiskott-Aldrich syndrome protein (WASP), and Bruton’s tyrosine kinase (BTK) in samples from patients with p91-phox deficiency, Wiskott-Aldrich syndrome (WAS), and X-linked agammaglobulinemia (XLA), respectively, were markedly lower than those in control samples (Fig. 3, E, H, and I). A sample from a patient with p22-phox deficiency also showed markedly reduced levels of its causative protein (Fig. 3 F). Interestingly, p22-phox levels paralleled those of p91-phox in this patient (Fig. 3, E and F), likely reflecting the tight interaction between these molecules (Parkos et al., 1989; Porter et al., 1994). A similar effect was observed in an FHL type 5 sample, in that Syntaxin-11 levels were reduced in parallel with those of Munc18-2 (Fig. 3, B and C). Levels of the causative protein were also lower in sample from a patient with p47-phox deficiency than those in healthy controls; however, they were only slightly below the normal range (Fig. 3 G). Samples from patients with Hermansky-Pudlak syndrome (HPS) type 2 also had reduced p47-phox protein levels (Fig. 3 G). By contrast, levels of lysosomal trafficking regulator (LYST), adaptor related protein complex 3 subunit beta 1 (AP3B1), and adenosine deaminase (ADA) in patients with Chédiak-Higashi syndrome (CHS), HPS type 2, and ADA deficiency, respectively, did not differ significantly from those in control samples (Fig. 3, J–L). Levels of common γ chain (IL2RG) were below the limit of detection in every sample. Absolute expression levels of the disease-causing protein in every patient, together with those in healthy controls, are summarized in Table 1.

**Figure 3.**
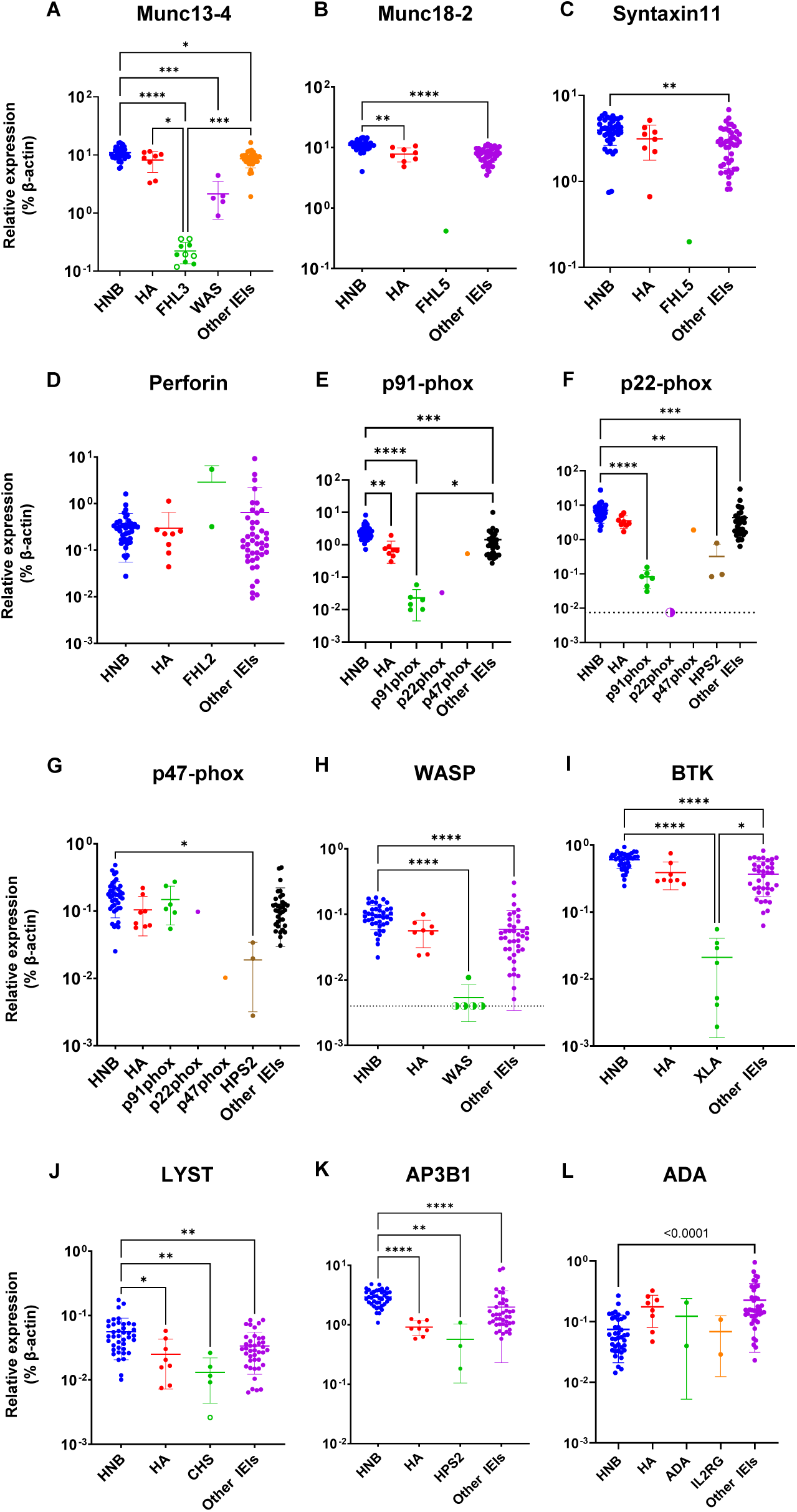
Expression of disease-responsible proteins in samples from patients with inborn errors of immunity (IEIs). Proteomics evaluation of disease-responsible proteins in samples from healthy newborns (HNB), healthy adults (HA), and patients with IEIs, including: familial hemophagocytic lymphohistiocytosis (FHL) type 2, 3, and 5 (FHL2: n = 2, FHL3: n = 8, and FLH5: n = 1, respectively); p91-phox, p22-phox, and p47-phox deficiencies (n = 6, 1, and 1, respectively); Hermansky-Pudlak syndrome type 2 (HPS2: n = 3); Wiskott-Aldrich syndrome (WAS: n = 5); X-linked agammaglobulinemia (XLA: n = 7); Chédiak-Higashi syndrome (CHS: n = 5); adenosine deaminase deficiency (ADA: n = 2); common γ chain deficiency (IL2RG: n = 2). All data were standardized relative to the expression of β-actin in the same sample. Levels of **(A)** Munc13-4, **(B)** Munc18-2, **(C)** Syntaxin-11, **(D)** perforin, **(E)** p91-phox, **(F)** p22-phox, **(G)** p47-phox, **(H)** WASP, **(I)** BTK, **(J)** LYST, **(K)** AP3B1, and **(L)** ADA in samples from HNB, HA, and patients with IEIs. Open symbols, samples obtained from pre-symptomatic newborn patients with IEIs; half-closed symbols, results under the limit of detection plotted at approximate lower detection limits. Statistical differences among groups were analyzed using one-way ANOVA on ranks (the Kruskal-Wallis test), followed by Dunn’s multiple comparisons. Data represent the mean ± SD. *\**P < 0.05, *\*\**P < 0.01, *\*\*\**P < 0.001, ****P < 0.0001.

We next determined whether the effect of a gene mutation on its protein level can be reliably predicted by *in silico* analysis. Table 2 shows the results of mutation effect predictions, together with the observed protein levels. As missense mutations often affect protein abundance by destabilizing protein structures (Yue et al., 2005), we performed computational prediction of the impact of missense mutations on protein stability using FoldX, a state-of-the art method for stability estimation based on the three-dimensional protein structures (Delgado et al., 2019). Of the 14 missense mutations identified in patients, 10 led to reduced protein abundance. Of those mutations leading to protein loss, 8 (80%) were successfully predicted to be destabilized, based on a total energy change threshold of > 1.58 kcal/mol for affecting protein stability upon a missense mutation (Gerasimavicius et al., 2020); however, two mutations, namely p.Pro359Thr in WASP and p.Arg525Gln in BTK, were falsely predicted to result in protein loss. Further, three mutations, namely p.Gly220Arg in perforin, p.Met63Lys in AP3B1, and p.Arg211His in ADA, were predicted to destabilize the proteins, while proteins harboring the mutations showed no loss of abundance in our experiments. These results indicate that use of an *in silico* approach is not sufficiently reliable, in terms of sensitivity and specificity, to evaluate the impact of missense mutations on protein abundance.

**Table 2.**
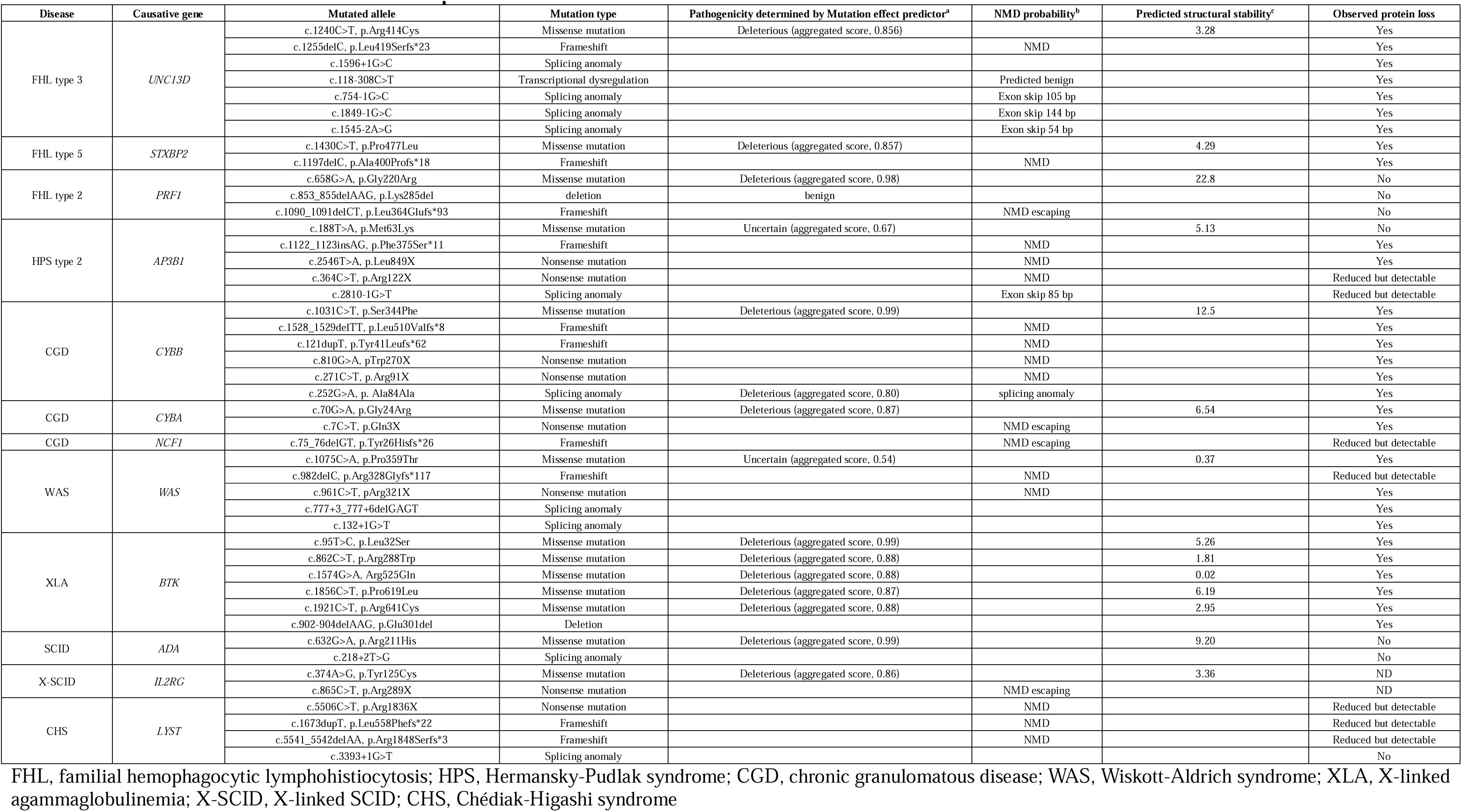
Predicted and observed levels of mutated proteins.

### Secondary effects of IEI gene mutations on DBS protein profiles

Next, we evaluated the secondary effects of IEI mutations and searched for proteins that could serve as alternative markers for diagnosis/screening of IEIs, particularly those that were unidentifiable by direct evaluation of the causative gene products. For accurate evaluation of DEPs between patients and healthy controls, age-related physiological alterations in protein levels must be taken into account; however, because of their extreme rarity, collection of newborn DBS samples for every IEI is almost impossible. Furthermore, secondary protein profile modification must be considered, since many patient samples were collected after disease onset. Therefore, we searched for possible disease markers by evaluating changes in expression beyond the level of age-dependent variation and secondary modification after disease onset. To this end, we first compared the protein profiles of DBS samples from patients with specific diseases with those from healthy newborns and with those from adults. We then searched for biological pathways that were commonly altered in patient samples relative to both control groups, followed by identification of specific molecules in these pathways that were also in the list of DEPs.

The neutrophil degranulation pathway was commonly altered in patients with HPS type 2 relative to healthy newborns and adults, as well as in patients with CHS compared to the two control groups (Fig. 4, A and B). Among common DEPs shared in this pathway, levels of p22-phox (*CYBA*), cathepsin G (*CTSG*), and myeloperoxidase (*MPO*), were reduced in samples from patients with HPS type 2, whereas those of cathepsin G and ELANE (*ELANE*) were reduced in patients with CHS relative to controls (Fig. 4 C). These findings likely reflect the phenotypes of each disease; that is, neutropenia in HPS type 2 and cell granule abnormalities in CHS. By combining ADA and common γ chain deficiencies together as SCID, immune system and neutrophil degranulation pathways were commonly altered in patients with SCID compared to healthy newborns and adults (Fig. 4 D). Among common DEPs in these pathways, levels of CD3E were exclusively lower in samples from patients with SCID than in healthy controls (Fig. 4 E), likely reflecting the phenotypic alteration in blood cell numbers in this disease (T cell lymphopenia). No protein was found to be common to pathways that were altered in patients with FHL type 2 relative to healthy newborns and adults.

**Figure 4.**
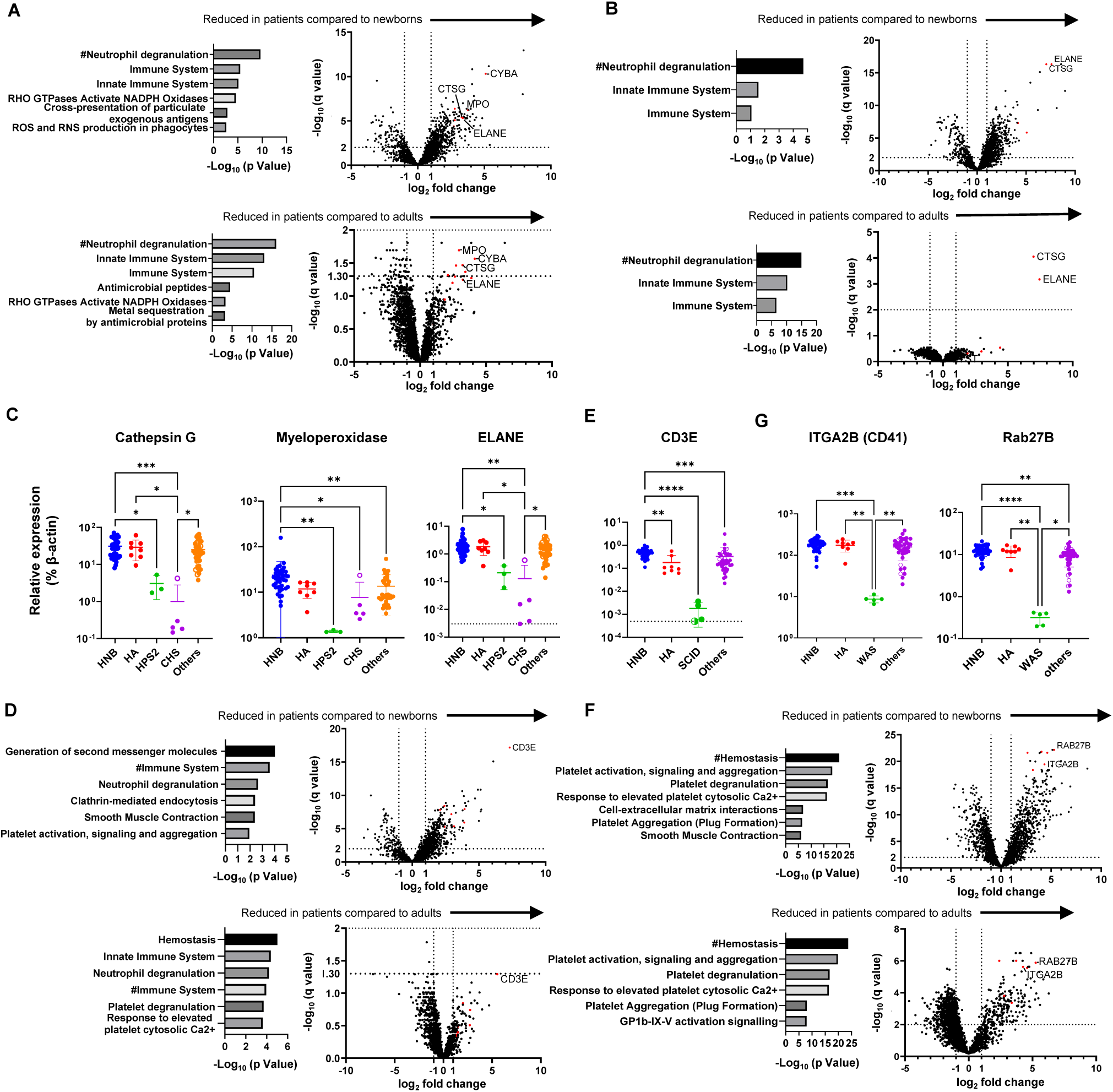
Identification of marker proteins associated with molecular phenotypes of inborn errors of immunity (IEIs). Gene ontology enrichment analysis of differentially expressed proteins (DEPs) (left) and volcano plots showing DEPs (right) in dried blood spot (DBS) samples from patients with **(A)** Hermansky-Pudlak syndrome type 2 (HPS2: n = 3), **(B)** Chédiak-Higashi syndrome (CHS: n = 5), **(D)** SCID: n = 4, and **(F)** Wiskott-Aldrich syndrome (WAS: n = 5) relative to samples from healthy newborns (HN) (left) and healthy adults (HA) (right). Levels of **(C)** cathepsin G, myeloperoxidase, and ELANE, **(E)** CD3E, and **(G)** ITGA2B (CD41) and Rab27B in DBS samples from HNB, HA, and patients with IEIs. Statistical differences among groups were analyzed using one-way ANOVA on ranks (the Kruskal-Wallis test), followed by Dunn’s multiple comparisons. Data represent the mean ± SD. *\**P < 0.05, *\*\**P < 0.01, *\*\*\**P < 0.001, ****P < 0.0001. #, Top Gene Ontology terms commonly altered in patient samples compared to both newborn and adult control groups. Red dots in volcano plots indicate proteins shared in biological pathways commonly altered in patient samples relative to those from HN and HA.

Interestingly, the hemostasis pathway was commonly altered in patients with WAS relative to healthy newborns and adults (Fig. 4 G). Among the common DEPs in this pathway, CD41 (*ITGA2B*) and Rab27b (*RAB27B*) were selectively reduced in patients with WAS, likely reflecting thrombocytopenia (Fig. 4 H). This explains the reduction of Munc13-4 levels in WAS samples, as Munc13-4 is abundant in platelets (Fig. 4 A) (Murata et al., 2011). We also found that levels of proteins related to T lymphocytes (CD2, CD3D, CD3E, CD5, CD247), platelets (CD41, CD42b), and neutrophils (CD11b, CD18, CD33, CD66b) were reduced in patients with T cell lymphopenia, thrombocytopenia, and neutropenia, respectively, and could be clustered as distinct groups (Fig. 5), suggesting that assays of these proteins could be used to identify patients with these cell-phenotypical alterations.

**Figure 5.**
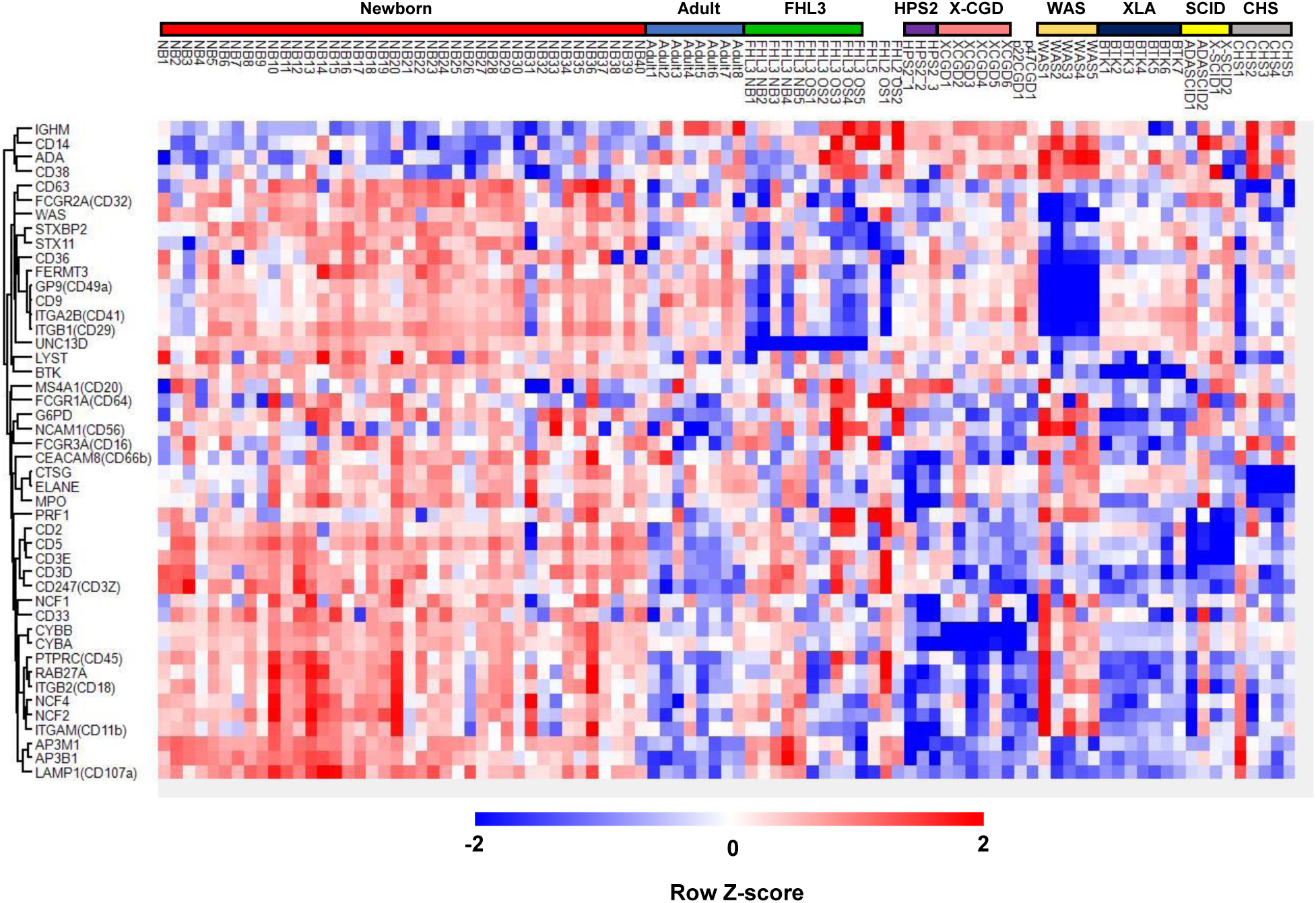
Comparison of proteomics data from patients with inborn errors of immunity with those from healthy newborns and adults. Heatmap showing hierarchical clustering of the levels of 42 proteins that are disease-responsive or associated with molecular phenotypes in dried blood spot samples from healthy newborns, healthy adults, and patients with inborn errors of immunity. Red, relatively higher expression; blue, relatively lower expression.

## Discussion

Screening for genetic diseases enables early identification of patients who need life-saving treatment before disease onset. The Wilson and Jungner recommendations for population-based screening have guided decisions regarding candidate disease inclusion in NBS programs for the past 50 years; however, the changing landscape of clinical practice and newer technologies, including genome-based methodologies, as well as developing perspectives on the ethical, legal, and social implications of population-based screening practices, necessitate review of these historic criteria (King et al., 2021). The analysis of multiple genes in newborns raises ethical considerations, and difficulties arise in interpreting variants with yet undetermined significance. The primary purpose of NBS is to convincingly detect significant signs of suspected genetic disease at the pre-symptomatic stage, rather than to conclusively diagnose newborns. Thus, it is reasonable that IEMs have been screened by measuring metabolite levels or activities of metabolic enzymes, as a first-tier test, as abnormal levels of the metabolite or enzymatic activity of interest are directly relevant to IEM pathogenesis; however, suitable markers for other inherited diseases, which can be used to detect functional anomalies at molecular and/or cellular levels before macroscopic disease signs manifest, remain elusive. In this context, we examined the applicability of quantitative protein profiling of DBS, supported by recent progress in DIA-LC-MS/MS technologies, including the development of a simple method for pre-treatment of DBS for non-targeted proteome measurements (Nakajima et al., 2020).

Using the newly-developed DIA-LC-MS/MS method, we successfully identified and quantified 2912 proteins in all healthy newborn samples tested. Evaluation of these proteins in newborn DBS samples revealed that their profiles were strongly correlated among newborn samples, which clustered distinctly from adult samples (Fig. 1, A and B). In agreement with previous reports, newborn DBS contained more fetus-specific proteins (AFP, fetal Hb-related components), as well as proteins related to translational initiation pathways and cell-cell adhesion, and fewer IgA/IgM-related proteins and proteins associated with coagulation and complement pathways (Fig. 2, A–D). Of consistently quantified proteins, 1110 were listed in OMIM and 781 were related to diseases with autosomal recessive or X-linked inheritance (Table S2). These included candidate diseases for NBS, such as Menkes disease, Wilson disease, congenital platelet abnormalities, and coagulation factor deficiencies, as well as various IEIs that have potential for screening by detection of reduced levels of disease-responsible proteins.

IEIs are caused by monogenic germline variants that result in loss or gain of function of the encoded proteins, and patients with these conditions present with increased susceptibility to a spectrum of infections, as well as various autoimmune, autoinflammatory, allergic, and/or malignant phenotypes (Bousfiha et al., 2022). IEIs are associated with significant morbidity and mortality if left untreated (King and Hammarström, 2018). Patients with severe IEIs require hematopoietic cell transplantation or other curative approaches, with earlier intervention resulting in better outcomes (Bakhtiar et al., 2021; Bergsten et al., 2020; Burroughs et al., 2020; Chiesa et al., 2020; Lucchini et al., 2018). Measuring T cell receptor excision circles (TRECs) and kappa-deleting element recombination circles (KRECs) in DBS samples can identify patients with T and B cell lymphopenias, respectively, and facilitates early treatment intervention and reduction of overall medical costs (Borte et al., 2012; Chan and Puck, 2005; Elsink et al., 2020; King et al., 2021; Nakagawa et al., 2011; Sheller et al., 2020). These results indicate that feasible NBS-based methods to detect other severe IEIs could improve patient outcomes. Furthermore, live vaccines administered in early infancy often cause opportunistic infections in certain groups of infants with IEIs, such as those with SCID and chronic granulomatous disease (Conti et al., 2016), which could be prevented by NBS.

Our data demonstrate that non-targeted DBS proteomics analysis can detect reduced levels of disease-responsible proteins in samples from patients with FHL type 3, FHL type 5, p91-phox, and p22-phox deficiencies, WAS, and XLA, relative to healthy newborn control samples (Fig. 3, A, B, E, F, H, and I). Moreover, the levels of p22-phox and Syntaxin-11 were reduced in parallel with those of p91-phox and Munc18-2, respectively (Fig. 3, B–C, E–F), in agreement with previous reports (Parkos et al., 1989; zur Stadt et al., 2009). Munc13-4 protein was specifically decreased in all pre-symptomatic newborn samples from patients with FHL type 3, which had been used for screening for metabolic disorders and stored for months to years, in the same way as detected in post-symptomatic samples (Fig. 3 A). These findings demonstrate the robustness of DBS proteomics for NBS of IEIs and also suggest its potential usefulness for diagnosing patients living in regions where other diagnostic modalities are unavailable.

The levels of perforin, LYST, and ADA proteins were not significantly reduced in patients with FHL type 2, CHS, and ADA deficiency, respectively (Fig. 3, D, J and L). Many *PRF1* mutations result in inappropriate folding or polymerization of perforin molecules, with residual protein expression on western blotting analysis (Voskoboinik et al., 2005). Further, many *LYST* mutations result in residual expression of truncated protein (Certain et al., 2000), which cannot be distinguished from full-length LYST on LC-MS/MS, unless a specific peptide near the C-terminus is evaluated. Furthermore, measurement of ADA by the current method may not be reliable because ADA is predominantly cytosolic and hydrophilic. Interestingly, AP3B1 levels varied greatly among patients with HPS type 2, with the highest level in a patient with a splicing site mutation near the 3’ end of one allele (Patient #13 in Table 1) and the lowest level in a patient who had experienced an episode of hemophagocytic lymphohistiocytosis (Patient #12 in Table 1) (Fig. 3 K), suggesting that HPS type 2 disease severity may correlate with residual AP3B1 expression.

As our method was non-targeted, we could also detect disease markers that reflect disease-associated phenotypes. Levels of p22-phox, cathepsin G, and myeloperoxidase were lower in DBS from patients with HPS type 2, and those of cathepsin G and ELANE were lower in patients with CHS, compared with controls (Fig. 4 C). These findings reflect the clinical phenotypes of the conditions; neutropenia in patients with HPS type 2 (Jung et al., 2006), and abnormal azurophil granules in patients with CHS (Burnett et al., 1995). As p22-phox, ELANE, and cathepsin G are primarily expressed in granulocytes, analyses of these proteins may also be useful in identifying patients with other diseases that are accompanied by neutropenia. CD3E levels were found to be reduced in patients with SCID (Fig. 4 E), and those of Rab27B and platelet-related proteins were low in patients with WAS (Fig. 4 H), likely reflecting thrombocytopenia and T cell lymphopenia, respectively, in these two IEIs. These results demonstrate that DBS proteomics can detect major changes in the number and/or characteristics of specific cell populations that can currently be analyzed only in specialized laboratories (Fig. 5). Notably, our method could identify patients with T cell lymphopenia and BTK deficiency and may complement TRECs/KRECs analyses, which have high sensitivity but relatively low specificity. DBS proteomics could be performed in parallel with TRECs/KRECs screening, using an additional punchout from the same sample, thus providing phenotypic specificity.

Another important aspect of the current study is that proteome analysis can provide information more directly relevant to patient conditions than that generated by genetic analysis. As shown in Table 2, the effects of genetic variants on levels of their encoded proteins are often unpredictable using available *in silico* analysis methods, and interpretation is difficult unless pathogenicity has been reported previously. Non-targeted DBS proteomics can provide in-depth molecular phenotype information and will aid in linking gene structural information with clinical phenotypes.

We acknowledge that this study has several limitations. First, most IEI samples analyzed here were obtained after disease onset, with only six pre-symptomatic neonatal samples included; five from patients with FHL type 3 and one from a patient with CHS. Therefore, further analysis of newborn IEI samples is required to prove the usefulness of the current method for NBS; however, most IEIs analyzed in this study are characterized by significant reductions in expression of disease-related proteins, or in specific cell populations, during the newborn period, which enables the diagnosis of siblings of index patients during their neonatal period before disease onset. In addition, healthy newborns experience physiological leukocytosis, resulting in high expression of leukocyte-related proteins during the neonatal period, and the proteins analyzed in the current study were consistently expressed in all healthy newborn samples. Thus, we do not anticipate that detecting reduced levels of disease marker proteins in newborn samples would be difficult. Second, not all patients display reduced marker protein levels, but protein expression levels often correlate with disease severity (Almarza Novoa et al., 2018; Kuhns et al., 2010; Zhu et al., 1997) and screening for patients with reduced protein levels would be of significant clinical impact. Third, we conducted DIA proteomic analysis with LC-MS/MS to evaluate a wide variety of non-target proteins, which is more time consuming than targeted analysis, such as monitoring of selected or parallel reactions, and could lead to challenges in providing prompt results for NBS; however, rapid advances in MS instrumentation and DIA-LC-MS/MS technology have led to the development of deep non-targeted proteome analysis with short measurement time (Ishikawa et al., 2022; Guzman et al., 2023). Therefore, many patients with IEIs characterized by significant reductions in disease marker proteins could be amenable to screening using non-targeted DIA-LC-MS/MS methods in the near future. Forth, we did not evaluate the unit cost of the test in the current study; the low incidence of severe genetic diseases means that the unit cost of a test is key to its potential for application. The method described in this study can be applied to a wide variety of congenital disorders, including metabolic/neurological and hematological diseases, associated with reduced levels of causative proteins in hydrophobic fractions of peripheral blood cells. Further, this method can readily be performed in parallel with other screening methods that use the same DBS samples, including TRECs/KRECs analysis and proteome analysis with pre-enrichment of target proteins using specific antibodies. Furthermore, advances in proteomics technology may expand the range of detectable peptides from disease-causing gene products, to allow detection of single amino acid alterations in abundant proteins (Salz et al., 2021). Together, such approaches will increase the number of disorders that can be screened simultaneously and reduce the per disease cost. In addition, DBS samples can be used for genome-wide genetic analysis, to confirm diagnoses and potentially lead to early intervention. Although additional improvements are needed for population-based applications, methodological refinements and innovations will enable the use of this approach to screen newborns for various diseases and strengthen the application of genome-wide sequencing-based screening in the future (King et al., 2021; Strand et al., 2020).

In conclusion, our findings demonstrate that levels of multiple proteins associated with genetic disorders in DBS samples can be semi-quantified, with no need for pre-enrichment using specific antibodies. Proteomic analysis of DBS samples can serve as a “single-platform-to-assess-many-diseases” NBS approach for IEIs and various other genetic disorders, enabling the identification of patients before disease onset, allowing earlier intervention in the absence of severe complications, and improving patient prognosis.

## Materials and methods

### Patients and samples

DBS samples on Guthrie cards from 40 healthy newborns used to screen for metabolic disorders were collected, as were samples from 8 healthy adults, and 45 samples from 43 patients with 12 IEI diseases (Table 1). Of the IEI samples, 38 were obtained after disease onset and stored at their respective institutions for research purposes, and the remaining FHL type 3 (n = 5) and CHS (n = 1) samples were obtained pre-symptomatically for use in public health screening for metabolic diseases. The study protocol, which conformed to the principles of the Declaration of Helsinki, was approved by the ethics committee of Kyoto University Hospital. Written informed consent was obtained from all participants or their legal guardians.

### Preparation and digestion of the insoluble protein fraction

Soluble fractions were reduced and proteins in DBS digested as previously described (Nakajima et al., 2020), with slight modification. Briefly, a disc (3.2 mm diameter) punched from a DBS was suspended in 1 mL of Na_2_CO_3_ solution and vigorously shaken in the presence of a 5 mm zirconia bead (Tomy Seiko, Tokyo, Japan) using a Tissue Lyser (Qiagen, Hilden, Germany) oscillating at 25 Hz for 5 min at room temperature. The remaining disc was removed by centrifugation (3,000 × g, 3 min, 4°C) and the supernatant (approximately 850 μL) was centrifuged (17,400 *g*, 15 min, 4°C). The precipitate was resuspended in 200 μL 100 mM sodium carbonate aqueous solution by vigorous vortexing at room temperature. After centrifugation (17,400 *g*, 5 min, 4°C), the supernatant was completely removed and the precipitate dissolved in 100 μL 12 mM sodium deoxycholate, 12 mM N-lauroylsarcosine, and 100 mM Tris-Cl pH 8.0 by vigorous vortexing for 1 min, followed by sonication using Bioruptor II (CosmoBio, Tokyo, Japan) on high power mode for 10 min in 30 s on/30 s off cycles. A 500 ng aliquot of trypsin/lys-C mix (Promega, Madison, WI) was added to each protein extract and samples were digested overnight at 37°C. After precipitation, detergent was removed by acidification with 30 μL 5% trifluoroacetic acid, followed by sonication with a Bioruptor II in high power mode for 5 min in 30 s on/30 s off cycles. The mixture was shaken for 5 min and centrifuged (15,000 *g*, 5 min). The supernatant was desalted using GL-Tip SDB (GL Sciences Inc., Tokyo, Japan), according to the manufacturer’s instructions, and dried using a centrifugal evaporator (miVac Duo concentrator, Genevac, Ipswich, UK). Dried peptides were redissolved in 3% ACN, 0.1% formic acid.

### LC-MS/MS

Digested peptides were directly injected onto a 75 μm × 20 cm PicoFrit emitter (New Objective, Woburn, MA) packed in-house with C18 core-shell particles (CAPCELL CORE MP 2.7 μm, 160 Å material; Osaka Soda Co., Ltd., Osaka, Japan) at 50°C and peptides separated using an UltiMate 3000 RSLCnano LC system (Thermo Fisher Scientific, Waltham, MA), with an 80 min gradient at 100 nL/min. Peptides eluted from the column were analyzed on an Orbitrap Exploris 480 Mass Spectrometer (Thermo Fisher Scientific) for overlapping window DIA-MS (Amodei et al., 2019; Kawashima et al., 2019). MS1 spectra were collected in the range, 495–785 m/z, at 30,000 resolution, to set an automatic gain control target of 3 × 10^6^ and maximum injection time of “auto”. MS2 spectra were collected in the range, 200–1800 m/z, at 30,000 resolution to set an automatic gain control target of 3 × 10^6^, maximum injection time of “auto”, and stepped normalized collision energies of 22%, 26%, and 30%. The MS2 isolation width was set to 4 m/z, and overlapping window patterns of 500–780 m/z used for window placements optimized by Skyline (20.2.0.34, University of Washington, Seattle, WA) (MacLean et al., 2010).

### MS data analysis

MS files were compared with human spectral libraries using Scaffold DIA (2.2.1, Proteome Software, Inc., Portland, OR). Human spectral libraries were generated from the human protein sequence database (UniProt id UP000005640, reviewed, canonical) using Prosit (https://www.proteomicsdb.org/prosit/) (Gessulat et al., 2019; Searle et al., 2020). Scaffold DIA search parameters were as follows: experimental data search enzyme, trypsin; maximum missed cleavage sites, 1; precursor mass tolerance, 9 ppm; and fragment mass tolerance, 9 ppm. The protein identification threshold was set at < 1% for both peptide and protein false discovery rates. Proteins and peptides were quantified using the EncyclopeDIA algorithm in Scaffold DIA (Searle et al., 2018).

### Prediction of effects of missense mutations on protein stability

Crystal structures of the proteins encoded by the disease-causing genes ADA, CYBA, CYBB, IL2RG, and STXBP2 were downloaded from Protein Data Bank (https://pdbj.org/) (PDB codes: 3iar for ADA; 8gz3 for CYBA and CYBB; 5m5e for IL2RG; 4cca for STXBP2). Where there were no experimentally determined structures for proteins with mutated residues (namely, AP3B1, PRF1, UNC13D, and WAS), protein model structures were obtained from the AlphaFold Protein Structure Database (https://alphafold.ebi.ac.uk/). Computational predictions of the impacts of missense mutations on protein stability were evaluated using FoldX, which is a state-of-the art method for stability estimation based on the three-dimensional protein structures (Delgado et al., 2019). If the total energy stability change value of the mutant protein was > 1.58 kcal/mol, the missense mutation was predicted to affect protein stability, following a previous report (Gerasimavicius et al., 2020).

### Statistical analysis

Protein expression levels were standardized using those of β-actin (%β-actin). Exploratory analysis, including log_2_ transformation, multiple scatter plotting, calculation of Pearson correlation coefficient values between two sets of data, data imputation (width = 0.3, Down shift = 2.4), Z-score normalization, and hierarchical comparison were performed using the Perseus software platform (https://maxquant.net/perseus/). Volcano plots were created by multiple t test using the two-stage step up method of Benjamini, Krieger, and Yekutieli in GraphPad Prism (version 9.0.2) software (GraphPad Software, Inc., La Jolla, CA). DEPs between two groups were selected for further analysis using a cutoff of adjusted P value (q value) < 0.01 and fold-change ≥ 2. Statistical differences among groups were analyzed using one-way ANOVA on ranks (Kruskal-Wallis test), followed by Dunn’s multiple comparisons test using GraphPad Prism. A P value < 0.05 was considered significant.

## Supplemental material

**Fig. S1** shows a Wikipathway map of the Complement and coagulation cascades highlighting downregulated proteins (P < 0.05) in healthy newborns relative to adults. **Fig. S2** illustrates quantification of proteins responsible for various genetic disorders in neonatal DBS samples. **Fig. S3** demonstrates that Munc13-4 is conspicuously reduced in DBS samples from patients with FHL type 3. **Fig. S4** presents flow cytometric analysis of perforin expression in natural killer cells from patients with familial hemophagocytic lymphohistiocytosis type 2. **Table S1** lists proteins identified in DBS samples analyzed in this study. **Table S2** lists proteins identified in DBS samples that are cataloged in the OMIM database. Table S1 and S2 are available at https://doi.org/10.5281/zenodo.10539971.

## Data availability

All of the data used to generate the figures are available in the published article and its online supplemental material.

## Acknowledgments

The authors are grateful to all participating patients and their families. This work was supported by JSPS KAKENHI under Grant Numbers 19K17328 and 21K07795; by AMED under Grant Number JP23ek0109586; and by the “Research on Measures for Intractable Diseases” project from the Japanese Ministry of Health, Labor, and Welfare.

## Authorship

Contributions: H.S., O.O., Y.K., and T.Y. designed the research; H.S., M.H., S.S., T. Miyamoto, and N-I.M. performed flow cytometric analyses of patients with FHL and collected their DBS samples; D.N., R.K., and Y.K. performed proteome analyses; K.O. and O.O. performed genetic analyses; A.H. and O.O. performed *in silico* prediction of missense mutation effects on protein stability; H.O. provided DBS samples from healthy babies; K. Izawa, H.O., M.I., M.Y., H.K., Y.N., S.N., T.U., M.O., and R.N. attended to patients and provided their samples; H.S., M.H., E.H., K. Izawa, J.T., T.H., K. Imai, T. Morio, O.O., Y.K., and T.Y. analyzed the data and discussed the results; and H.S., Y.K., and T.Y. wrote the paper.

## Correspondence

Takahiro Yasumi, Department of Pediatrics, Kyoto University Graduate School of Medicine, 54 Shogoin-Kawahara-cho, Sakyo-ku, Kyoto, 606-8507 JAPAN; Yusuke Kawashima, Kazusa DNA Research Institute, 2-5-23 Kazusa-Kamatari, Kisarazu, 292-0818, JAPAN

## Abbreviations

CHS: Chédiak-Higashi syndrome
DBS: dried blood spot
DEPs: differentially expressed proteins
DIA: data-independent acquisition
FHL: familial hemophagocytic lymphohistiocytosis
GO: Gene Ontology
HPS: Hermansky-Pudlak syndrome
IEIs: inborn errors of immunity
IEMs: inborn errors of metabolism
KRECs: kappa-deleting element recombination circles
LC-MS/MS: liquid chromatography-assisted mass spectrometry
NBS: newborn screening
OMIM: Online Mendelian Inheritance in Man
TRECs: T cell receptor excision circles
WAS: Wiskott-Aldrich syndrome
XLA: X-linked agammaglobulinemia

## Supplementary figure legends

**Figure S1.**
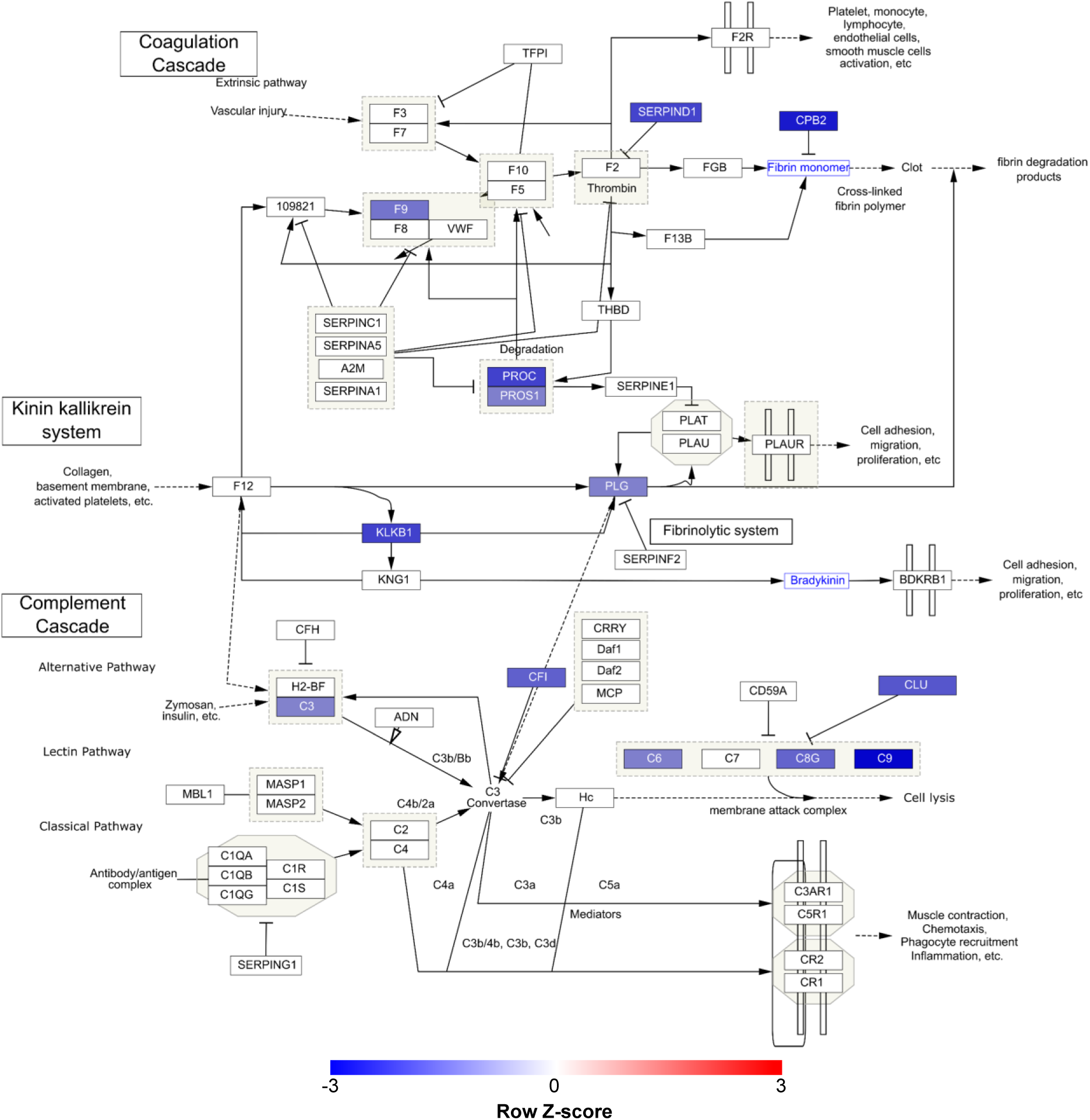
Wikipathway map of the Complement and coagulation cascades. Pathways were retrieved and visualized from WikiPathways using Cytoscape, highlighting downregulated proteins (P < 0.05) in healthy newborns relative to adults. Blue node colors denote differentially expressed proteins and fold changes in their expression. Blue characters indicate metabolite molecules.

**Figure S2.**
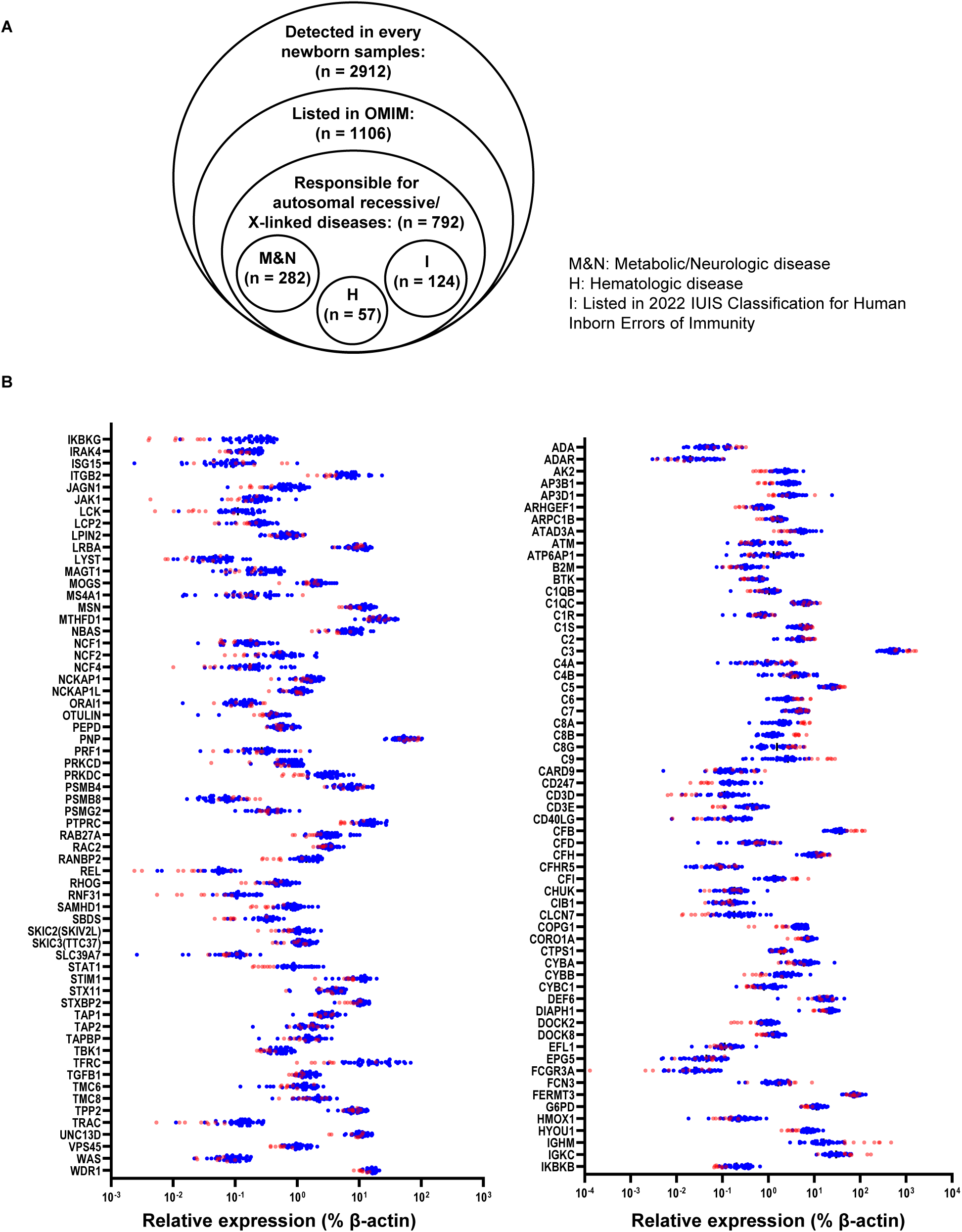
Quantification of proteins responsible for various genetic disorders in neonatal dried blood spot (DBS) samples. **(A)** Venn diagram of proteins identified in DBS samples. OMIM, Online Mendelian Inheritance in Man. **(B)** Levels of 124 proteins listed in the 2022 International Union of Immunological Societies classifications of inborn errors of immunity in DBS samples from healthy newborns (blue) and healthy adults (red).

**Figure S3.**
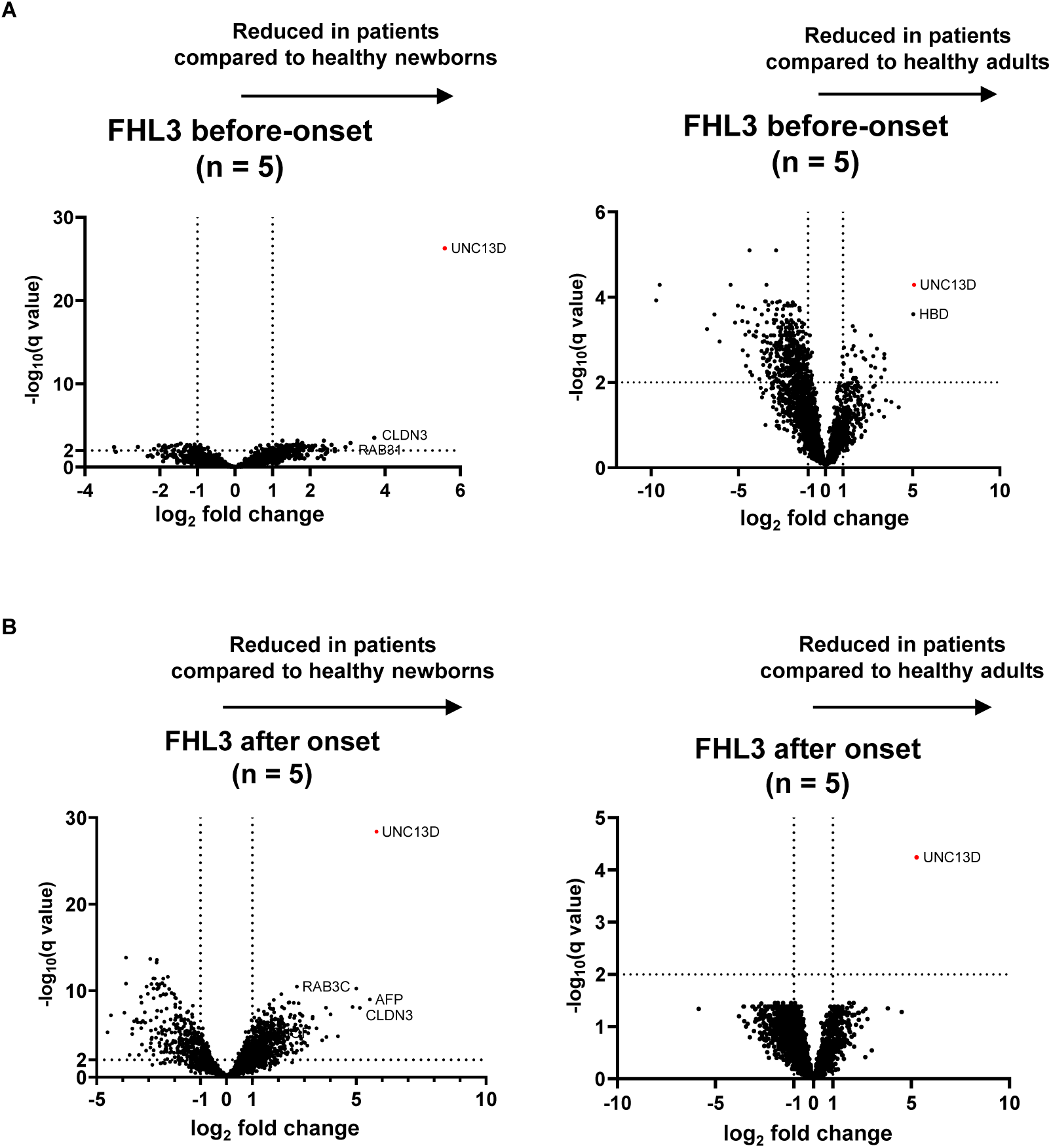
Munc13-4 is conspicuously reduced in dried blood spot (DBS) samples from patients with familial hemophagocytic lymphohistiocytosis type 3 (FHL3). Volcano plots showing the differentially expressed proteins in DBS samples from patients with FHL3 that were obtained **(A)** before and **(B)** after disease onset compared to samples from healthy newborns (left) and from healthy adults (right).

**Figure S4.**
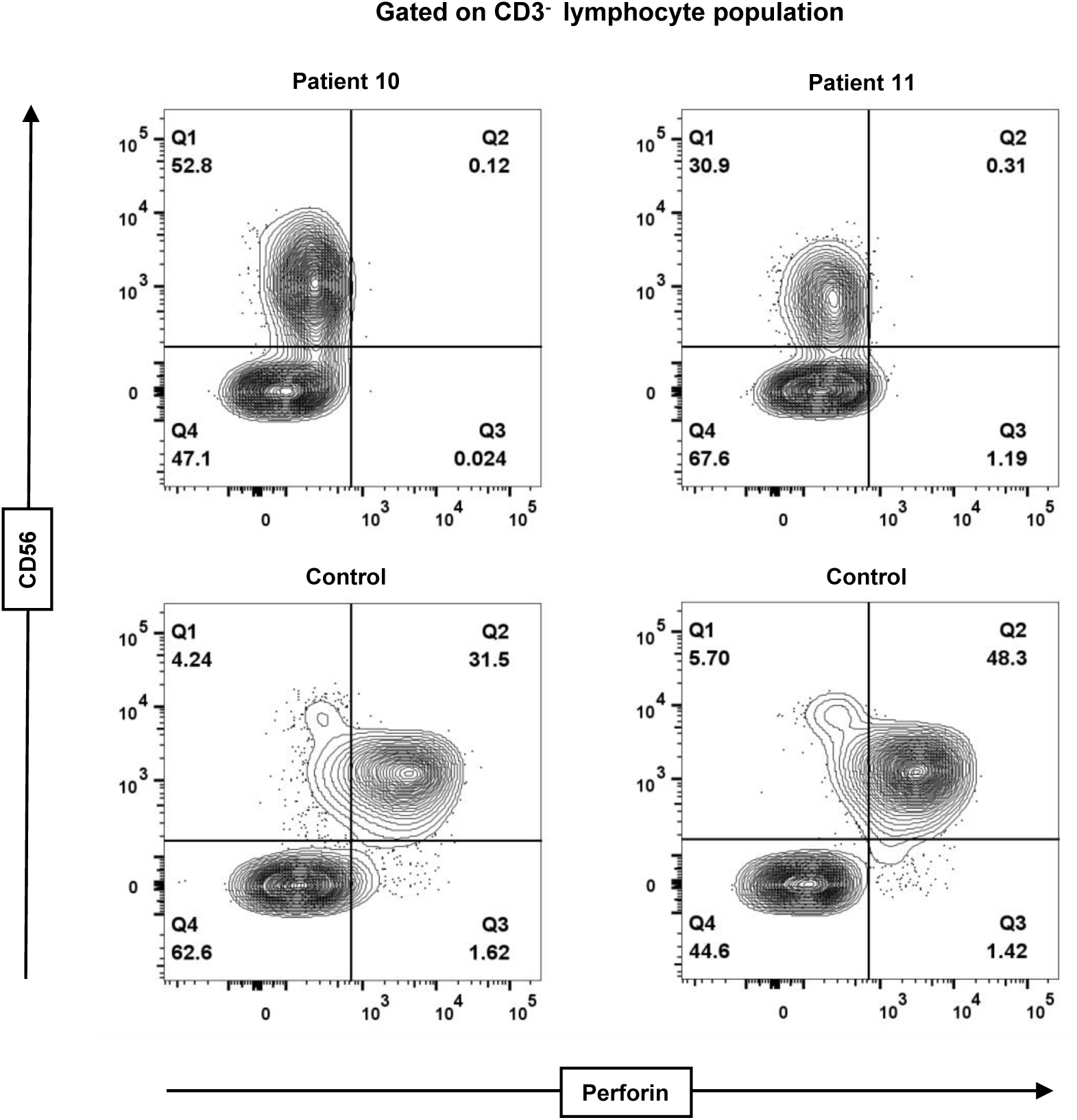
Flow cytometric analysis of perforin expression in natural killer cells from patients with familial hemophagocytic lymphohistiocytosis type 2. Fluorescence-activated cytometric analysis of perforin expression in CD3^-^ peripheral lymphocytes obtained from patients 10 and 11 relative to healthy controls.

## Supplemental table legends

**Table S1. List of proteins identified in dried blood spot samples analyzed in this study.**

The list includes identified proteins with peptide and protein false discovery rates < 1%. Proteins shown in black are those identified by unique peptide sequences that were clearly distinguishable from others. Proteins shown in blue represent grouped master proteins. Proteins shown in grey represent grouped proteins. A quantitative value of “0” indicates that the protein was detected, but did not reach a level sufficient for quantification.

**Table S2. Proteins identified in dried blood spot samples that are listed in the Online Mendelian Inheritance in Man (OMIM) database.**

List of proteins identified in every healthy newborn sample that are included in the OMIM database. The list includes identified proteins with peptide and protein false discovery rates < 1%. AD, autosomal dominant; AR, autosomal recessive; ARLOF, autosomal recessive loss of function; XL, X-linked; IUIS2022, 2022 Classification of Human Inborn Errors of Immunity by the International Union of Immunological Societies Expert Committee.

